# Large Language Models for Detecting CONSORT Guideline Compliance in Published Randomized Clinical Trials: A Cross-Sectional Evaluation Study

**DOI:** 10.1101/2025.10.03.25337291

**Authors:** Daniel Y Tsybulnik, Justin J Gillette, Thomas F Heston

## Abstract

**Background:** Peer review processes may inadequately assess compliance with established reporting guidelines such as the Consolidated Standards of Reporting Trials (CONSORT) criteria. Large language models (LLMs) demonstrate potential for systematic manuscript evaluation; however, their accuracy in detecting adherence to CONSORT guidelines in published clinical trials remains unexplored.

**Methods:** This cross-sectional study evaluated the compliance of 20 randomized controlled trials published between 2015 and 2024 from immunology journals, identified through PubMed, with the CONSORT 2010 guidelines. Three large language models (ChatGPT-4o, Gemini 2.5 Pro, and Claude Sonnet 4) independently assessed compliance across 37 CONSORT subpoints. The primary endpoint was the mean CONSORT compliance percentage. Secondary endpoints included the proportion of articles meeting a 90% compliance threshold and agreement between LLM assessments. Statistical analysis employed repeated measures ANOVA with post-hoc pairwise comparisons (α = 0.05).

**Results:** Mean CONSORT compliance rates were: ChatGPT-4o 81% (95% CI: 77-85%), Claude Sonnet 4 68% (95% CI: 61-75%), and Gemini 2.5 Pro 55% (95% CI: 48-62%). Overall compliance across all LLMs was 68% (95% CI: 64-72%). Using a 90% compliance threshold as a quality benchmark, ChatGPT-4o identified 25% of papers (5/20), Claude Sonnet 4 identified 5% (1/20), and Gemini 2.5 Pro identified none (0/20) as meeting this standard. Repeated-measures ANOVA demonstrated significant differences in LLM performance (F_2,38_ = 40.79, p < 0.001, partial η^2^ = 0.682). All pairwise comparisons between models were statistically significant (p ≤ 0.002).

**Conclusions:** Large language models detected CONSORT compliance deficiencies in published randomized trials, aligning with previously reported rates of 60-70%, which validates their accuracy in identifying persistent reporting quality issues. The substantial variation between LLM assessments indicates the need for standardized evaluation protocols. These findings support the potential utility of LLM-assisted manuscript evaluation to improve adherence to established reporting guidelines.

## Introduction

Peer review serves as the primary quality control mechanism for scientific publications, yet studies consistently demonstrate substantial limitations in detecting methodological errors and reporting deficiencies (1). The increasing strain on peer review processes, driven by rapid growth in manuscript submissions, has affected evaluation rigor across publishers (2). When investigators intentionally introduced nine significant errors into randomized controlled trial manuscripts, peer reviewers detected an average of only three errors, with nearly 25% of reviewers identifying one error or fewer (3). These systematic limitations extend to harm data reporting, where inconsistent analysis in randomized controlled trials fails to provide adequate information for clinical decision-making (4).

The Consolidated Standards of Reporting Trials (CONSORT) provides an evidence-based, minimum set of recommendations for reporting the methodology and results of randomized controlled trials to ensure clarity and transparency. First published in 1996 and subsequently updated in 2001, 2010, and 2025, CONSORT guidelines facilitate critical appraisal and enhance research reproducibility (5). Despite widespread endorsement in high-impact medical journals, compliance remains suboptimal. Analysis of 463 abstracts from five leading journals found overall adherence of 67%, with individual journal rates ranging from 55% to 78% (6).

Large language models (LLMs) have increasingly been utilized in scholarly processes, including literature reviews, manuscript preparation, and systematic error detection (7). Advanced models, such as GPT-4, detect approximately 53% of intentionally inserted errors, approaching the performance of human peer reviewers (8). When comparing LLM feedback to human reviewer comments on 3,096 manuscripts from Nature family journals, the overlap between GPT-4 and individual human reviewers (31%) closely matched inter-human reviewer agreement (29%) (9). The practical significance of these capabilities was demonstrated when an AI model identified, within seconds, a ten-fold mathematical error in published flame-retardant research—a miscalculation that human reviewers had missed. This detection prompted the Black Spatula Project, an open-source initiative employing LLMs to identify overlooked errors in scientific literature (10). LLMs have also demonstrated utility in automated paper screening for clinical reviews, extending their application to systematic literature evaluation (11).

However, the ability of LLMs to evaluate compliance with structured reporting guidelines such as CONSORT remains unexplored. Given persistent suboptimal adherence to CONSORT guidelines and documented limitations of human peer review in detecting reporting deficiencies, automated assessment of guideline compliance represents a critical gap in quality control mechanisms. This investigation evaluated the accuracy of three leading LLM platforms in detecting CONSORT guideline adherence in published randomized controlled trials.

## Methods

### Study Design and Objectives

This cross-sectional investigation evaluated compliance of randomized clinical trials published between 2015 and 2024 with CONSORT 2010 guidelines. The temporal restriction to pre-2025 publications ensured consistency with the 2010 criteria, as the CONSORT guidelines were updated in April 2025 (5).

### Study Selection and Inclusion Criteria

Randomized controlled trials were identified through a PubMed search using the search strategy: randomized controlled trial [Publication Type] AND immunology. The first 20 trials meeting the inclusion criteria were selected for evaluation. Trials published after the 2025 CONSORT update were excluded to maintain methodological consistency throughout the evaluation period.

### Sample Size Determination

This study employed an adaptive design approach to optimize statistical efficiency while maintaining methodological rigor. Data collection proceeded sequentially, with interim analysis conducted after 20 randomized controlled trials to assess whether adequate statistical power had been achieved. The decision to terminate data collection at N=20 was based on: (1) achievement of statistical significance with substantial effect sizes between LLM assessments, (2) post-hoc power analysis using G*Power 3.1.9.7 confirming excellent statistical power (81.7%) to detect meaningful differences in CONSORT compliance assessment between the three large language models using a repeated measures design with α = 0.05, and (3) resource optimization principles consistent with adaptive trial methodology. The large observed effect size (partial η^2^ = 0.811) validated that sufficient data had been collected to draw definitive conclusions regarding LLM performance differences.

### CONSORT Evaluation Framework

The CONSORT 2010 checklist encompasses 25 primary items subdivided into 37 specific subpoints, each addressing essential elements of trial methodology and reporting (12). This framework provides standardized criteria for evaluating the quality and transparency of randomized controlled trial reporting.

### Scoring Methodology

Each article was evaluated across all 37 CONSORT subpoints using a three-level scoring system: complete fulfillment (1.0 point), partial fulfillment (0.5 points), or non-fulfillment (0 points). The large language models independently determined fulfillment levels based on whether each CONSORT criterion was fully addressed with complete detail as specified in the CONSORT 2010 guidelines (1.0 point), partially addressed with incomplete detail or clarity (0.5 points), or not addressed (0 points). Overall compliance was calculated as the proportion of maximum possible points achieved across all subpoints.

### CONSORT Compliance Threshold

A 90% compliance threshold was selected as a clinically meaningful benchmark for high-quality reporting. While current adherence rates in leading medical journals range from 55% to 78%, with overall rates of approximately 67% (6), the CONSORT guidelines represent minimum standards for transparent trial reporting. For journals that endorse CONSORT guidelines and publish research frequently cited in clinical practice, near-complete adherence to minimum reporting standards represents a reasonable quality expectation. This threshold serves as an aspirational benchmark against which current reporting practices can be evaluated.

### Large Language Model Implementation

Three large language model platforms conducted parallel evaluations: ChatGPT-4o (OpenAI), Gemini 2.5 Pro (Google), and Claude Sonnet 4 (Anthropic). ChatGPT-4o and Gemini 2.5 Pro received standardized prompts with detailed instructions for criterion-by-criterion analysis and justifications. Claude Sonnet 4 was evaluated using a simplified binary assessment prompt to investigate whether the complexity of the prompt affects evaluation consistency. Complete PDF manuscripts were uploaded to each platform, generating one independent evaluation per article per model. Complete prompts provided to each LLM platform are available in Supplement A. The list of evaluated articles with DOIs is provided in Supplement B.

### Quality Control and Validation

All evaluated articles had previously undergone traditional peer review and been accepted for publication in immunology journals indexed in PubMed that require adherence to CONSORT guidelines for randomized trial reporting. The LLM-detected mean CONSORT compliance of 68% (95% CI: 64-72%) aligns with previously reported compliance rates of 60-70% in published randomized trials (13), validating model accuracy in detecting known reporting deficiencies.

### Statistical Analysis

Statistical analyses were conducted following SAMPL (Statistical Analyses and Methods in the Published Literature) guidelines. A repeated measures ANOVA was performed with an α level of 0.05. The primary endpoint was CONSORT compliance percentage across 37 subpoints. Secondary endpoints included the proportion of articles meeting a 90% compliance threshold and inter-model agreement in assessments. Descriptive statistics included means and 95% confidence intervals. Post-hoc pairwise comparisons were performed using the Bonferroni correction for multiple testing. Effect sizes are reported as partial eta-squared (η^2^). Statistical analyses were conducted using IBM SPSS Statistics version 31 (IBM Corp., Armonk, NY, USA).

### Ethical Considerations

This investigation utilized exclusively publicly available, peer-reviewed journal articles. No human subject research was performed.

## Results

### CONSORT Compliance Assessment

The three large language models demonstrated substantial variability in CONSORT compliance assessment. Mean compliance rates were: ChatGPT-4o 81% (95% CI: 77-85%), Claude Sonnet 4 68% (95% CI: 61-75%), and Gemini 2.5 Pro 55% (95% CI: 48-62%). The overall compliance rate across all three LLMs was 68% (95% CI: 64-72%).

### Quality Threshold Analysis

Using a 90% compliance threshold as a quality benchmark, ChatGPT-4o identified 5 articles (25%), Claude Sonnet 4 identified 1 article (5%), and Gemini 2.5 Pro identified no articles (0%) as meeting this standard. Conversely, 75-100% of papers failed to meet the 90% threshold, depending on the LLM evaluator. These findings indicate substantial deficiencies in CONSORT reporting standards even among peer-reviewed publications (Figure 1).

**Figure 1.**
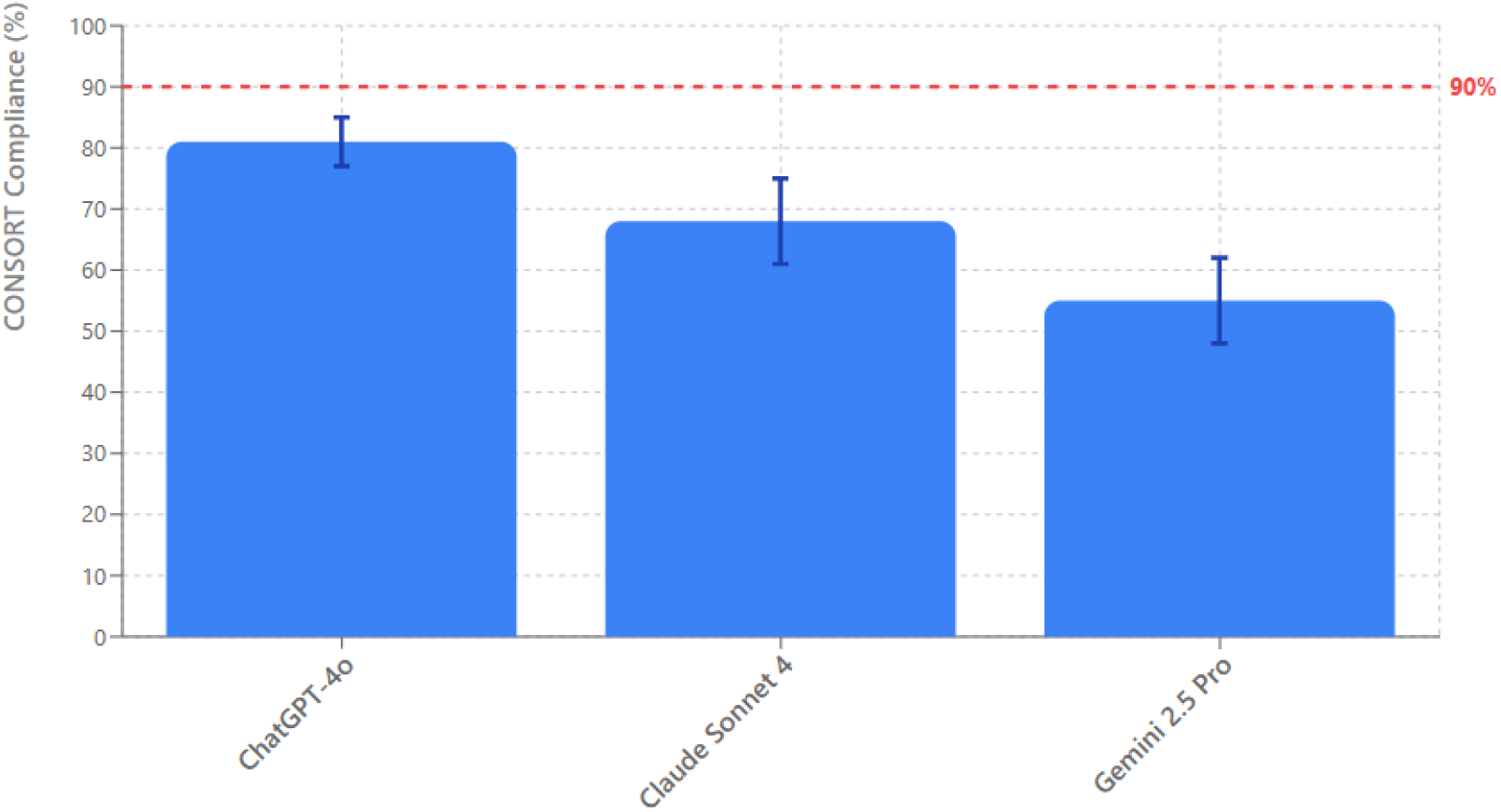
CONSORT Compliance Assessment by Large Language Model Platform. *Note*. Mean CONSORT 2010 compliance rates for 20 randomized controlled trials evaluated by three large language model platforms. Error bars represent 95% confidence intervals. The dashed line at 90% indicates the quality threshold benchmark. N=20 articles per platform. Compliance was assessed using a three-level scoring system (0, 0.5, 1.0 points) across 37 CONSORT 2010 subpoints. Statistical analysis revealed significant differences between all pairwise comparisons (p ≤ 0.002 for all comparisons, repeated measures ANOVA F_2,38_ = 40.79, p < 0.001).

### Statistical Comparison of LLM Performance

Repeated-measures ANOVA demonstrated a statistically significant main effect of LLM type on CONSORT compliance scores (F_2,38_ = 40.79, p < 0.001, partial η^2^ = 0.682). This large effect size indicates substantial differences in how the three models assessed compliance.

Post-hoc pairwise comparisons using Bonferroni correction revealed statistically significant differences for all comparisons: ChatGPT-4o versus Claude Sonnet 4 (p = 0.001), ChatGPT-4o versus Gemini 2.5 Pro (p < 0.001), and Claude Sonnet 4 versus Gemini 2.5 Pro (p = 0.002). ChatGPT-4o demonstrated the highest compliance detection rates, followed by Claude Sonnet 4, then Gemini 2.5 Pro.

### Power Analysis

Post-hoc power analysis using G*Power 3.1.9.7 confirmed excellent statistical power (>99%) for detecting the observed differences between LLM evaluators, validating the adequacy of the sample size for addressing the research objectives.

## Discussion

This investigation demonstrates that large language models can detect CONSORT compliance deficiencies in published randomized controlled trials, with an overall compliance rate of 68% that aligns closely with previously reported rates of 60-70% in the literature (13). However, the study reveals substantial variation between LLM platforms, with ChatGPT-4o demonstrating the most lenient assessment (81% mean compliance), Claude Sonnet 4 showing intermediate stringency (68% compliance), and Gemini 2.5 Pro applying the most stringent criteria (55% compliance). Notably, even the most optimistic LLM assessment found that only 25% of papers met the 90% compliance threshold expected for high-quality clinical journals.

These findings occur within the broader context of well-documented peer review limitations (1). Previous investigations demonstrate that peer reviewers detect only 3 of 9 intentionally inserted significant errors in randomized controlled trial manuscripts, with 25% of reviewers identifying only one error and 16% failing to detect any errors at all (3). The increasing strain on peer review processes, driven by rapid growth in manuscript submissions, has reduced the time available for thorough evaluation of research quality and methodology (2). Different peer review procedures demonstrate varying abilities to flag problematic publications, with systematic limitations across traditional editorial processes (14). This investigation suggests that LLMs may serve as valuable complementary tools to address these systematic limitations in conventional editorial processes.

The alignment between LLM-detected compliance rates and established literature provides essential validation of model accuracy in identifying reporting deficiencies. This validation is particularly significant given documented examples of LLM success in detecting errors missed by peer review, such as the ten-fold dosage miscalculation in brominated diethyl ether research that prompted the Black Spatula Project—a grassroots initiative using LLMs to identify overlooked academic errors (10). Previous studies consistently report CONSORT compliance rates between 60-70% across medical specialties, suggesting that the observed 68% overall rate reflects genuine reporting limitations rather than systematic LLM over-detection (15). LLMs have demonstrated capabilities in detecting commonly missed errors, including lack of protocols, ambiguous ethics statements, and inappropriate citation practices (16).

The finding that 75-100% of papers reviewed in this study failed to meet a 90% CONSORT compliance rate based on LLM review represents a clinically significant concern for evidence-based medicine. Inadequate CONSORT compliance compromises the ability of clinicians and researchers to appraise study methodology critically, assess risk of bias, and apply findings to patient care (17). The potential for missed reporting errors to affect real-world clinical decisions underscores the importance of systematic quality assessment tools. Clinical research errors that escape detection can have substantial consequences, as evidenced by high-profile retractions due to statistical analysis problems, such as the 2023 retraction of a highly cited medication adherence study due to misleading statistical reporting (18). Among all reasons why a medical article may be deemed unfit for publication, the most common are plagiarism and data fabrication (19), but broader applications for error detection remain underutilized.

The substantial differences between LLM assessments indicate that model architecture, training data, and prompt design significantly influence evaluation outcomes (7). ChatGPT-4o’s more lenient scoring may reflect training on a broader corpus of published literature, potentially normalizing suboptimal reporting practices. Conversely, Gemini 2.5 Pro’s stringent assessment may indicate more precise adherence to CONSORT criteria specifications. Notably, when comparing LLM feedback to human reviewer comments on 3,096 papers from Nature family journals, the overlap between LLM and human assessments (31%) closely matched inter-human reviewer agreement (29%), with 82.4% of users finding GPT-4 feedback more helpful than some human reviewers (9). However, recent comparative studies show that while LLMs demonstrate reliable internal consistency, they differ substantially from human reviewers in manuscript evaluation, with humans rejecting 68.2% of manuscripts compared to LLM rejection rates of 0-9.1% (20).

LLMs demonstrate particular strengths in linguistic editing, summarization, and systematic guideline evaluation, but may show limitations with statistics-heavy or nuanced medical content (7). Previous research indicates that while LLMs can identify various types of errors comparable to human reviewers, they may exhibit weaknesses in evaluating complex statistical analyses and mathematical content (8). Some studies have raised concerns about LLM limitations, including the generation of confident-sounding hallucinations with little relationship to actual paper content (21). With certain prompting techniques or retrieval-augmented generation, newer LLM iterations can achieve lower hallucination rates on specific tasks compared to earlier models, though these improvements are often limited and do not eliminate the underlying issue (22). Natural language processing approaches show promise in addressing medically inaccurate information and various forms of research errors (20).

These results extend previous investigations, demonstrating that GPT-4 detected 53% of intentionally inserted errors, comparable to the performance of human peer reviewers (8). However, the current investigation focuses on systematic guideline compliance rather than error detection, revealing that LLMs can identify patterns of reporting deficiencies across published literature (23). This represents a novel application distinct from previous work on linguistic editing, summarization, and basic error detection, addressing a critical gap in evaluating the quality of clinical trial reporting, which serves as the backbone of medical knowledge advancement. Studies have demonstrated ChatGPT’s capability in identifying methodological flaws and providing insightful feedback on theoretical frameworks, with critical analyses aligning with human reviewers (24). Recent work has also demonstrated LLM effectiveness in automated paper screening for clinical reviews, extending their utility beyond individual manuscript assessment to systematic literature evaluation (11).

Several limitations affect the interpretation of these findings. First, this study evaluated only immunology journals, which may limit generalizability; however, a single-specialty focus enabled controlled assessment without confounding. Second, the different prompt designs between platforms may explain some of the performance differences, although this study explored whether prompt complexity affects consistency. The partial scoring system (0/0.5/1.0) introduced subjective interpretation; conversely, it reflects real-world evaluation where criteria are often partially met. Third, no gold-standard human expert assessment was conducted. While the 68% compliance aligns with literature rates of 60-70%, validating consistency, absolute accuracy remains unestablished. Nevertheless, external validation against benchmarks suggests LLMs identify known reporting deficiencies. Fourth, the adaptive design, while statistically justified, was not pre-specified; however, post-hoc power analysis and large effect size validate sample sufficiency. Fifth, LLMs may exhibit limitations in statistical content (7), although CONSORT predominantly assesses reporting transparency. Finally, CONSORT compliance represents only one quality dimension; however, this focused scope provides clear evidence for LLM capability in structured guideline evaluation.

Future research should focus on developing standardized LLM evaluation protocols for manuscript assessment, conducting head-to-head comparisons with expert human reviewers, and expanding evaluation to additional reporting guidelines such as PRISMA and STARD. Investigation of prompt engineering techniques to optimize LLM performance and reduce inter-model variability represents another critical research priority. Additionally, exploration of LLM capabilities for detecting other forms of research misconduct, including statistical fraud and citation manipulation, warrants investigation, given the documented limitations of traditional peer review in identifying such issues (14). The continued evolution of LLM capabilities, with each model release showing improved efficiency and accuracy, suggests expanding potential applications in scientific quality assessment (25).

## Conclusions

Large language models demonstrate substantial capability for detecting CONSORT compliance deficiencies in published clinical trials, with accuracy validated by alignment with established literature and documented peer review limitations. However, significant variability between LLM platforms emphasizes the need for standardized evaluation protocols before widespread implementation in editorial processes. These findings support the potential utility of LLM-assisted manuscript evaluation as a complementary tool for enhancing adherence to established reporting guidelines, addressing well-documented limitations in traditional peer review processes, and highlighting the persistent challenge of ensuring high-quality reporting in clinical trials.

## Supporting information

Supplement A - LLM Prompts

Supplement B - Evaluated Articles List

## Data Availability Statement

Raw data supporting the findings of this study are openly available in Zenodo at https://doi.org/10.5281/zenodo.17253371

