## Supplement A - LLM Prompts for "Large Language Models for Detecting CONSORT Guideline Compliance in Published Randomized Clinical Trials: A Cross-Sectional Evaluation Study"

Specific prompts for ChatGPT-4o, Gemini 2.5, and Claude Sonnet 4

### Prompt for ChatGPT -4o and Gemini 2.5

You are an expert published reviewer. Read the attached research paper thoroughly, ensuring complete understanding of its theoretical foundations, methodology, statistical analyses, data interpretations, conclusions and every other characteristic of the paper.

Then thoroughly familiarize yourself with the 2010 CONSORT statement which is used to improve the quality of reporting of RCTs, and which consists of a checklist and flow diagram that authors can use for reporting an RCT.

CONSORT 2010 checklist of information to include when reporting a randomised trial*

Title and abstract

1a Identification as a randomised trial in the title

1b Structured summary of trial design, methods, results, and conclusions (for specific guidance see CONSORT for abstracts)

Introduction

Background and objectives

2a Scientific background and explanation of rationale

2b Specific objectives or hypotheses

Methods

Trial design

3a Description of trial design (such as parallel, factorial) including allocation ratio

3b Important changes to methods after trial commencement (such as eligibility criteria), with reasons

Participants

4a Eligibility criteria for participants

4b Settings and locations where the data were collected

Interventions

5 The interventions for each group with sufficient details to allow replication, including how and when they were actually administered

Outcomes

6a Completely defined pre-specified primary and secondary outcome measures, including how and when they were assessed

6b Any changes to trial outcomes after the trial commenced, with reasons

Sample size

7a How sample size was determined

7b When applicable, explanation of any interim analyses and stopping guidelines

Randomisation

Sequence generation

8a Method used to generate the random allocation sequence

8b Type of randomisation; details of any restriction (such as blocking and block size)

Allocation concealment mechanism

9 Mechanism used to implement the random allocation sequence (such as sequentially numbered containers), describing any steps taken to conceal the sequence until interventions were assigned

Implementation

10 Who generated the random allocation sequence, who enrolled participants, and who assigned participants to interventions

Blinding

11a If done, who was blinded after assignment to interventions (for example, participants, care providers, those assessing outcomes) and how

11b If relevant, description of the similarity of interventions

Statistical methods

12a Statistical methods used to compare groups for primary and secondary outcomes

12b Methods for additional analyses, such as subgroup analyses and adjusted analyses

Results

Participant flow (a diagram is strongly recommended)

13a For each group, the numbers of participants who were randomly assigned, received intended treatment, and were analysed for the primary outcome

13b For each group, losses and exclusions after randomisation, together with reasons

Recruitment

14a Dates defining the periods of recruitment and follow-up

14b Why the trial ended or was stopped

Baseline data

15 A table showing baseline demographic and clinical characteristics for each group

Numbers analysed

16 For each group, number of participants (denominator) included in each analysis and whether the analysis was by original assigned groups

Outcomes and estimation

17a For each primary and secondary outcome, results for each group, and the estimated effect size and its precision (such as 95% confidence interval)

17b For binary outcomes, presentation of both absolute and relative effect sizes is recommended

Ancillary analyses

18 Results of any other analyses performed, including subgroup analyses and adjusted analyses, distinguishing pre-specified from exploratory

Harms

19 All important harms or unintended effects in each group (for specific guidance see CONSORT for harms)

Discussion

Limitations

20 Trial limitations, addressing sources of potential bias, imprecision, and, if relevant, multiplicity of analyses

Generalisability

21 Generalisability (external validity, applicability) of the trial findings

Interpretation

22 Interpretation consistent with results, balancing benefits and harms, and considering other relevant evidence

Other information

Registration

23 Registration number and name of trial registry

Protocol

24 Where the full trial protocol can be accessed, if available

Funding

25 Sources of funding and other support (such as supply of drugs), role of funders

You will be evaluating randomized control trial articles in regards to how well the article follows the checklist.

Next, I want a checklist that clearly specifies whether or not the paper met each of the 37 subpoints of the CONSORT criteria. I also want you to evaluate for two outcome variables: how well the paper conforms to the CONSORT criteria by percentage (how much of the subpoints were met over the total of all the subpoints) as well as the severity of deviation from the CONSORT criteria. In regards to the severity of deviation from the Consort criteria, I want a low severity label as a count of 0-3 items that were not or were only partially met, a medium label as 4-7 items that were not or were only partially met and a high severity label as a count of 8 or more items that were not or were only partially met. No table please. For each item that was partially met or not met, I want you to give an explanation of how exactly it did not meet the subpoint.

### Claude Sonnet 4

USER: I am a physician peer-reviewer of medical research.

GPT ROLE: you are to act as an expert review assistant that meticulously analyzes each manuscript given to you for compliance with the 2010 CONSORT criteria.

2010 CONSORT CRITERIA:

**Title and Abstract**

- 1a: Identification as a randomised trial in the title

- 1b: Structured summary of trial design, methods, results, and conclusions

**Introduction**

- 2a: Scientific background and explanation of rationale

- 2b: Specific objectives or hypotheses

**Methods**

- 3a: Description of trial design (such as parallel, factorial) including allocation ratio

- 3b: Important changes to methods after trial commencement (such as eligibility criteria), with reasons

- 4a: Eligibility criteria for participants

- 4b: Settings and locations where the data were collected

- 5: The interventions for each group with sufficient details to allow replication, including how and when they were actually administered

- 6a: Completely defined pre-specified primary and secondary outcome measures, including how and when they were assessed

- 6b: Any changes to trial outcomes after the trial commenced, with reasons

- 7a: How sample size was determined

- 7b: When applicable, explanation of any interim analyses and stopping guidelines

**Randomisation**

- 8a: Method used to generate the random allocation sequence

- 8b: Type of randomisation; details of any restriction (such as blocking and block size)

- 9: Mechanism used to implement the random allocation sequence (such as sequentially numbered containers), describing any steps taken to conceal the sequence until interventions were assigned

- 10: Who generated the random allocation sequence, who enrolled participants, and who assigned participants to interventions

- 11a: If done, who was blinded after assignment to interventions (for example, participants, care providers, those assessing outcomes) and how

- 11b: If relevant, description of the similarity of interventions

**Statistical Methods**

- 12a: Statistical methods used to compare groups for primary and secondary outcomes

- 12b: Methods for additional analyses, such as subgroup analyses and adjusted analyses

**Results**

- 13a: For each group, the numbers of participants who were randomly assigned, received intended treatment, and were analysed for the primary outcome

- 13b: For each group, losses and exclusions after randomisation, together with reasons

- 14a: Dates defining the periods of recruitment and follow-up

- 14b: Why the trial ended or was stopped

- 15: A table showing baseline demographic and clinical characteristics for each group

- 16: For each group, number of participants (denominator) included in each analysis and whether the analysis was by original assigned groups

- 17a: For each primary and secondary outcome, results for each group, and the estimated effect size and its precision (such as 95% confidence interval)

- 17b: For binary outcomes, presentation of both absolute and relative effect sizes is recommended

**Ancillary Analyses**

- 18: Results of any other analyses performed, including subgroup analyses and adjusted analyses, distinguishing pre-specified from exploratory

**Harms**

- 19: All important harms or unintended effects in each group

**Discussion**

- 20: Trial limitations, addressing sources of potential bias, imprecision, and, if relevant, multiplicity of analyses

- 21: Generalisability (external validity, applicability) of the trial findings

- 22: Interpretation consistent with results, balancing benefits and harms, and considering other relevant evidence

**Other Information**

- 23: Registration number and name of trial registry

- 24: Where the full trial protocol can be accessed, if available

- 25: Sources of funding and other support (such as supply of drugs), role of funders

GPT RESPONSE FORMAT

Reply with a table and nothing else.

Setup the table as follows:

- row 1: header row listing all of the CONSORT criteria as noted above

- row 2: list whether or not the criteria was met using the following categories: "Yes", "No", or "Partially". Use those responses verbatim. No other responses are allowed.

FOLLOW TABLE FORMATTING PRECISELY: 37 COLUMNS, 2 ROWS AS DESCRIBED ABOVE. THE TABLE MUST BE EASY TO CUT-N-PASTE THE RESULTS INTO A GOOGLE SHEET.

FOLLOW TABLE FORMATTING PRECISELY: 37 COLUMNS, 2 ROWS AS DESCRIBED ABOVE. THE TABLE MUST BE EASY TO CUT-N-PASTE THE RESULTS INTO A GOOGLE SHEET.

FOLLOW TABLE FORMATTING PRECISELY: 37 COLUMNS, 2 ROWS AS DESCRIBED ABOVE. THE TABLE MUST BE EASY TO CUT-N-PASTE THE RESULTS INTO A GOOGLE SHEET.
